# KNOWLEDGE AND PERCEPTION ABOUT HUMAN PAPILLOMAVIRUS IN TRADITIONAL QUILOMBOLA COMMUNITIES IN NORTHEAST BRAZIL

**DOI:** 10.1101/2024.10.11.24315317

**Authors:** Mariel Rodrigues de Campos, Larissa Helena Sousa Baldez Carvalho, Ana Julya Rodrigues Campos, Gabriel Rodrigues Côra, Rayane Alves Machado, Nafisa M.K Elehamer, Luisa Lina Villa, José de Ribamar Ross, Flávia Castello Branco Vidal

## Abstract

The study analyzed knowledge about Human Papillomavirus among adults in traditional quilombola communities in northeast Brazil while discussing their sociodemographic characteristics and evaluating sociodemographic variables among different quilombola communities. It was a descriptive, cross-sectional, and quantitative mixed-method study, with face-to-face interviews and a questionnaire application. There is a significant gap in the precise understanding of HPV. Most information was acquired through traditional media, such as TV, newspapers, and radio. Women and younger individuals demonstrated more comprehensive knowledge of the subject. The results highlight the need for specific instructions to improve knowledge about HPV in these communities, with strategies adapted to their sociodemographic characteristics. Enhanced understanding of HPV and the importance of vaccination can significantly contribute to the health and well-being of these populations, decreasing health disparities. With better knowledge about HPV and its consequences, the vaccination rate will increase, which in the long term will mean a reduction in the incidence of cancer cases associated with this virus, especially cervical cancer.

## INTRODUCTION

Human papillomavirus (HPV) is a DNA virus responsible for one of the most common sexually transmitted infections worldwide. High-risk HPV types are a cause of several important human cancers, especially in the genital area (Doorbar et al., 2012). According to WHO, cervical cancer (CC) was the fourth cause of women’s deaths until 2020, with the highest rate of CC incidence and mortality in low and middle-income countries (World Health statistics, 2023). HPV vaccination has reduced CC incidence in higher-income countries by up to 80%, although, in low and middle-income countries, there are still many obstacles to reaching this baseline (García et al., 2023). HPV vaccine is part of the annual vaccination program in various countries and can be applied in one or two doses in girls and boys between the ages of 9 and 14 before they initiate their sexual life (Alsous et al., 2021; Doorbar et al., 2012).. However, vaccination coverage depends on different factors, including access, cost, organization, and structure of health programs for awareness of the vaccine (Chandeying; Thongseiratch, 2023). Poor and low knowledge of the efficacy, security, and existence of HPV vaccine is still a barrier to HPV vaccination, especially in low and middle-income countries with low Human Development Index (HDI), conservative culture, and poor education (Waller et al., 2013; Gallagher et al., 2018).

Studies have been carried out all over the world to demonstrate the low knowledge of certain populations about HPV, its consequences, and prevention methods. Alsous and co-authors (2021)(Alsous et al., 2021) showed that only 26.1% of Arab women knew about HPV vaccination. Raçi and colleagues (2021) observed that 66.4% of Kosovo’s women had never heard about HPV (Raçi et al., 2021). Warner and co-authors (2022) evaluated the knowledge and awareness of HPV and HPV vaccination among an indigenous Caribbean community, showing that few had heard of HPV (19.6%) or the HPV vaccine (21.8%) (Warner et al., 2022). Even within underdeveloped countries, there are specific populations that are in even more vulnerable conditions that make them more predisposed to diseases caused by poor access to health care, especially sexual health. This is the case for indigenous people and other traditional communities around the globe (Warner et al., 2022;de Ribamar Ross et al., 2023). Quilombolas^1^ is a conventional Brazilian community composed of individuals descended from Africans enslaved between the 16th and 19th centuries. Some still live culturally and geographically isolated, having their form of social organization and reproduction of cultural habits (Dias et al., 2021).

Influential factors of HPV vaccination adherence include knowledge and perceived risk of HPV disease, misinformation about vaccine safety, and erroneous ideas like induction of sexual promiscuity. Public understanding of what HPV is and the forms of prevention is important to ensure the population’s participation in prevention programs(Waller et al., 2013). Studies aimed at measuring and monitoring public awareness and knowledge are important to guide health education campaigns to lead to correct knowledge and consequently, enhance HPV vaccination in quilombola and other traditional public [4-6]. Alsous and colleagues (2021) used a questionnaire via Google Forms sent through various media platforms to measure HPV knowledge among Arab women (Alsous et al., 2021). In Kosovo women, a questionnaire was distributed among women and self-answered(Raçi et al., 2021). In a systematic review and meta-analysis, Chandeying and Thongseiratch (2023) discussed interventional methods to improve HPV vaccination, especially in low-developed regions where the coverage of HPV vaccination is only 2.7%. They demonstrated that incorporating digital interventions such as reminders and alerts in mobile phones and social media platforms can effectively promote HPV vaccination (Chandeying; Thongseiratch, 2023).

In our study, we hypothesize that there is little knowledge about HPV, HPV vaccination, and prevention among quilombola populations. We expect similar results to researchers with the same evaluation in traditional communities with around 20% knowledge about HPV. We will include not only females but also quilombola males. We will carry out the questionnaires in person, reducing possible misunderstanding biases, despite taking longer and being more labour-intensive. Analysing knowledge about the HPV vaccine in traditional communities is essential to characterise the profile and scale of public health problems related to the issue (Warner et al., 2022).

## METHODS

### STUDY SETTINGS AND POPULATION

Maranhão is the second Brazilian state with the largest quilombola communities (IBGE, 2023). Caxias is a city in the extreme east of Maranhão with 21 quilombolas communities registered until June 2020 throughout the municipality’s territory (Lima et al., 2021). According to two studies on quilombola communities of Caxias, each quilombo has an average of 200 inhabitants; the sample calculation was based on the estimate of 1200 quilombola people, which provides a minimum number of 292 research participants (Lima et al., 2021; de Ribamar Ross et al., 2023). The calculation was done using the software https://www.surveymonkey.com/mp/sample-size-calculator/. This study will include six quilombola communities from Caxias named (1) Cajueiro, (2) Mimoso, (3) Soledade, (4) Lavras, (5) Jenipapo, and (6) Lagoa dos Pretos. The Brazilian map with the location of Maranhão state, the city of Caxias, and the studied quilombola communities are available in the supplementary material with the experimental methodology design.

### STUDY TYPE, PARTICIPANT RECRUITMENT, AND DATA COLLECTION

This cross-sectional survey included males and females aged ≥ 18 years, residents in the quilombola communities who declare themselves quilombolas. Exclusion criteria are those less than 18 years old. The questionnaire utilized to collect data was developed by the authors based on the scientific literature on the topic (Alsous et al., 2021; Raçi et al., 2021). The first section contains questions about general demographic data such as age, gender, quilombo of origin, occupation, level of education, family income, and marital status. The second section included questions to evaluate the general knowledge and awareness about HPV and its vaccine. What is HPV, its transmission, its correlation to genital warts and cervical cancer, prevention, HPV vaccine existence, availability, and if they would let their kids be vaccinated. Interviews were carried out individually with the participants and the researchers. The surveys were filled using a pen by trained research team members based on the participants’ oral responses. The survey responses were manually entered into Excel.

### STATISTICS

The answers obtained in the interviews were recorded on the SurveyMonkey® platform. The data resulting from these responses were subsequently tabulated using Microsoft Office Excel® (2016 version) from Microsoft Corporation (Redmond, WA, USA). Statistical analysis was conducted using the Statistical Package for the Social Sciences - SPSS (version 21), developed by IBM Corporation (Chicago, IL, USA). The demographic characteristics of the interviewees were described using descriptive statistics. Data presentation was performed using absolute (n) and relative frequency (%). To compare categorical variables between gender, age groups, and quilombo of residence, the Chi-Square or Fisher’s Exact (£) tests were applied, depending on the distribution. All statistical associations were set at a significance level of p ≤ 0.05.

### ETHICS

The study is approved by the Fundação Universidade Federal do Maranhão Research Ethics Committee, number: 5.836.549.

## RESULTS

### PARTICIPANTS’ SOCIODEMOGRAPHIC CHARACTERISTICS

A total of 302 individuals participated in the study. According to Table 01 of the sociodemographic characterisation of the sample, the average age was 41.42 years old, with the majority being women (61.3%), aged between 18 and 34 years old (38.4%). The Jenipapo community had the highest number of representatives in the survey (22.2%). The sample was mainly composed of farmers (28.8%). Education among the quilombola people was low, with a quarter of them declaring themselves illiterate and 29.1% having only primary education. The predominant marital status was married/stable union (50.3%) and a monthly income of up to a minimum wage (approximately $271.59). There was a diversity in the religious affiliations of the participating individuals, with a predominance of 59.9% identified as Catholic (Table 1). Regarding skin colour, the majority declared themselves black.

**Table 1.** Participants’ demographic data of 302 individuals from six quilombola communities in Caxias, Brazil.

| Variables | N (%) |
| --- | --- |
| **Median age ± SD (years)** | 41.42 ± 15.16 |
| **Stratified age (years)** |  |
| 18 - 34 | 116 (38.4%) |
| 35 - 49 | 91 (30%) |
| 50 - 64 | 70 (23.2%) |
| + 65 | 25 (8.3%) |
| **Sex** |  |
| Female | 185 (61.2%) |
| Male | 117 (38.8%) |
| **Occupation** |  |
| Farmer | 107 (35.4%) |
| Student | 53 (17.5%) |
| Others (guard, secretary) | 51 (16.9%) |
| Housewife | 36 (11.9%) |
| Unemployed | 30 (9.9%) |
| Retired | 25 (8.3%) |
| **Marital status** |  |
| Married | 152 (50.3%) |
| Divorced/Widow | 68 (22.5%) |
| Single | 82 (27.1%) |
| **Religion** |  |
| Catholic | 181 (59.9) |
| Evangelic | 79 (26.2) |
| African-based | 17 (5.6) |
| Others | 25 (8.3) |
| **Quilombola Community** |  |
| Jenipapo | 67 (22.2%) |
| Lavras | 51 (16.9%) |
| Soledade | 51 (16.9%) |
| Mimoso | 48 (15.9%) |
| Lagoa dos Pretos | 45 (14.9%) |
| Cajueiro | 40 (13.2%) |
| **Educational level** |  |
| Illiterate | 78 (25.8%) |
| Primary school | 88 (29.1%) |
| Secondary school | 103 (34.1%) |
| Undergraduate study | 33 (10.9%) |
| **Mensual family income (US Dollar)** |  |
| No income | 104 (34.4%) |
| Up to a minimum wage (271,59) | 145 (48%) |
| More than a minimum wage | 53 (17.6%) |
| **Self-declared color** |  |
| Yellow | 3 (1) |
| White | 50 (16.6) |
| Brown | 102 (33.8) |
| Black | 147 (48.7) |

### PARTICIPANTS’ KNOWLEDGE AND AWARENESS ABOUT HPV AND THE VACCINE

Knowledge and awareness about HPV and its vaccine were measured using an assisted questionnaire with closed questions of yes, no or I don’t know answers. (Table 2). Overall, both women and men reported having heard about HPV (88.1% and 86.3%). However, the knowledge rate decreased when asked what the type of microorganism was (55.7% and 54.7%), and the majority got the question wrong when reporting that HPV was a bacteria (86.5% and 82.9%). The main vehicle of informative communication about HPV was traditional media (TV/newspaper/radio) for both women (39.5%) and men (28.2%). The internet ranked 4th among women and 2nd among men. The majority reported knowing how it is transmitted (62.7% and 66.7%); they answered the question correctly that said whether transmission was sexual (70.8% and 76.9%), and both sexes knew that condoms prevent infection (64.9% and 70.9%).

**Table 2.** Knowledge and perceptions about HPV and its vaccination among men and women living in quilombola communities in Caxias, MA. São Luís, Maranhão, 2023

| HPV Questionnaire | Gender |  | Value of p ¥ |
| --- | --- | --- | --- |
|  | Female<br>n (%) | Male<br>n (%) |  |
| Have you heard of HPV? |  |  |  |
| Yes | 163 (88,1) | 101 (86,3) | 0,649 |
| No | 22 (11,9) | 16 (13,7) |  |
| You know what it is? |  |  |  |
| Yes | 103 (55,7) | 64 (54,7) | 0,868 |
| No | 82 (44,3) | 53 (45,3) |  |
| Is it a virus, bacteria or fungi? |  |  |  |
| Bacteria | 160 (86,5) | 97 (82,9) | 0,395 |
| Fungi | 17 (9,2) | 17 (14,5) |  |
| Virus | 107 (57,8) | 57 (48,7) |  |
| Where did you hear about it? |  |  |  |
| Friends | 6 (3,2) | 9 (7,7) | 0,159 |
| Internet/social media | 25 (13,5) | 24 (20,5) |  |
| Haven't heard | 22 (11,9) | 16 (13,7) |  |
| Teachers | 27 (14,6) | 15 (12,8) |  |
| Health professionals | 32 (17,3) | 20 (17,1) |  |
| TV/newspaper/radio | 73 (39,5) | 33 (28,2) |  |
| Do you know what genital warts are? (cockscomb)? |  |  |  |
| Yes | 82 (44,3) | 36 (30,8) | **0,019\*** |
| No | 103 (55,7) | 81 (69,2) |  |
| Is there a vaccine for HPV? |  |  |  |
| Yes | 120 (64,9) | 75 (64,1) | 0,187 |
| No | 13 (7) | 3 (2,6) |  |
| Do not know | 52 (28,1) | 39 (33,3) |  |
| Is there a vaccine at the nearby health facility? |  |  |  |
| Yes | 29 (15,7) | 17 (14,5) | 0,388 |
| No | 53 (28,6) | 26 (22,2) |  |
| Do not know | 103 (55,7) | 74 (63,2) |  |
| Would you vaccinate your child? |  |  |  |
| Yes | 165 (89,2) | 102 (87,2) | 0,595 |
| No | 20 (10,8) | 15 (12,8) |  |
| Do you know how HPV is transmitted? |  |  |  |
| Yes | 116 (62,7) | 78 (66,7) | 0,484 |
| No | 69 (37,3) | 39 (33,3) |  |
| Can it be transmitted through sexual intercourse? |  |  |  |
| Yes | 131 (70,8) | 90 (76,9) | 0,238 |
| No | 28 (15,1) | 18 (15,4) |  |
| Do not know | 26 (14,1) | 9 (7,7) |  |
| Do condoms prevent it? |  |  |  |
| Yes | 120 (64,9) | 83 (70,9) | 0,547 |
| No | 30 (16,2) | 16 (13,7) |  |
| Do not know | 35 (18,9) | 18 (15,4) |  |
| Do you have a vaccinated child? |  |  |  |
| Yes | 72 (38,9) | 25 (21,4) | **<0,001\*** |
| No | 110 (59,5) | 70 (59,8) |  |
| Do not know | 3 (1,6) | 22 (18,8) |  |
| Have you already been vaccinated? |  |  |  |
| Yes | 25 (13,5) | 9 (7,7) | 0,119 |
| No | 160 (86,5) | 108 (92,3) |  |
| Is there a cure for HPV? |  |  |  |
| Yes | 116 (62,7) | 69 (59) | 0,792 |
| No | 36 (19,5) | 26 (22,2) |  |
| Do not know | 33 (17,8) | 22 (18,8) |  |

¥: Chi-square; HPV: Human papillomavirus.

**TABLE 3.** Knowledge and perceptions about HPV and its vaccination among age groups of residents of quilombola communities in Caxias, MA. Best time for São Luís 2023 in Maranhão - Dates.

| HPV Questionnaire | Age (years) |  |  |  | p Value ¥ |
| --- | --- | --- | --- | --- | --- |
|  | 18 - 34 | 35 - 49 | 50 - 64 | 65 + |  |
|  | n (%) | n (%) | n (%) | n (%) |  |
| Have you heard of HPV? |  |  |  |  |  |
| Yes | 112 (96,6) | 83 (91,2) | 57 (81,4) | 12 (48,0) | <0,001 |
| No | 4 (3,4) | 8 (8,8) | 13 (18,6) | 13 (52,0) |  |
| You know what it is? |  |  |  |  |  |
| Yes | 80 (69) | 56 (61,5) | 27 (38,6) | 4 (16,0) | <0,001 |
| No | 36 (31) | 35 (38,5) | 43 (61,4) | 21 (84,0) |  |
| Is it a virus, bacteria or fungi? |  |  |  |  |  |
| Bacteria | 105 (90,5) | 76 (83,5) | 57 (81,4) | 19 (76,0) | 0,155 |
| Fungi | 6 (5,2) | 9 (9,9) | 10 (14,3) | 9 (36,0) |  |
| Virus | 84 (72,4) | 39 (42,9) | 34 (48,6) | 7 (28,0) |  |
| Where did you hear about it? |  |  |  |  |  |
| Friends | 5 (4,3) | 2 (2,2) | 5 (7,1) | 3 (12,0) | <0,001 £ |
| Internet/social media | 31 (26,7) | 15 (16,5) | 3 (4,3) | 0 (0,0) |  |
| Haven't heard | 4 (3,4) | 8 (8,8) | 13 (18,6) | 13 (52,0) |  |
| Teachers | 26 (22,4) | 11 (12,1) | 4 (5,7) | 1 (4,0) |  |
| Health professionals | 25 (21,6) | 15 (16,5) | 9 (12,9) | 3 (12,0) |  |
| TV/newspaper/radio | 25 (21,6) | 40 (44) | 36 (51,4) | 5 (20,0) |  |
| Do you know what genital warts are? |  |  |  |  |  |
| Yes | 39 (33,6) | 41 (45,1) | 31 (44,3) | 7 (28,0) | 0,179 |
| No | 77 (66,4) | 50 (54,9) | 39 (55,7) | 18 (72,0) |  |
| Is there a vaccine? |  |  |  |  |  |
| Yes | 91 (78,4) | 64 (70,3) | 31 (44,3) | 9 (36,0) | <0,001 £ |
| No | 5 (4,3) | 4 (4,4) | 5 (7,1) | 2 (8,0) |  |
| Do not know | 20 (17,2) | 23 (25,3) | 34 (48,6) | 14 (56,0) |  |
| Is there a vaccine at the nearby health facility? |  |  |  |  |  |
| Yes | 16 (13,8) | 16 (17,6) | 9 (12,9) | 5 (20,0) | 0,484 |
| No | 36 (31) | 24 (26,4) | 16 (22,9) | 3 (12,0) |  |
| Do not know | 64 (55,2) | 51 (56) | 45 (64,3) | 17 (68,0) |  |
| Would you vaccinate your child? |  |  |  |  |  |
| Yes | 108 (93,1) | 78 (85,7) | 60 (85,7) | 21 (84,0) | 0,250 |
| No | 8 (6,9) | 13 (14,3) | 10 (14,3) | 4 (16,0) |  |
| Do you know how it's transmitted? |  |  |  |  |  |
| Yes | 89 (76,7) | 69 (75,8) | 31 (44,3) | 5 (20,0) | **<0,001 £** |
| No | 27 (23,3) | 22 (24,2) | 39 (55,7) | 20 (80,0) |  |
| Can it be transmitted through sexual intercourse? |  |  |  |  |  |
| Yes | 92 (79,3) | 70 (76,9) | 47 (67,1) | 12 (48,0) | 0,029 |
| No | 15 (12,9) | 13 (14,3) | 12 (17,1) | 6 (24,0) |  |
| Do not know | 9 (7,8) | 8 (8,8) | 11 (15,7) | 7 (28,0) |  |
| Do condoms prevent it? |  |  |  |  |  |
| Yes | 86 (74,1) | 62 (68,1) | 43 (61,4) | 12 (48,0) | 0,008 |
| No | 21 (18,1) | 11 (12,1) | 8 (11,4) | 6 (24,0) |  |
| Do not know | 9 (7,8) | 18 (19,8) | 19 (27,1) | 7 (28,0) |  |
| Have a vaccinated child? |  |  |  |  |  |
| Yes | 36 (31) | 40 (44) | 15 (21,4) | 6 (24,0) | **<0,001 £** |
| No | 80 (69) | 45 (49,5) | 41 (58,6) | 14 (56,0) |  |
| Do not know | 0 (0) | 6 (6,6) | 14 (20) | 5 (20,0) |  |
| You have already been vaccinated? |  |  |  |  |  |
| Yes | 31 (26,7) | 1 (1,1) | 2 (2,9) | 0 (0,0) | **<0,001 £** |
| No | 85 (73,3) | 90 (98,9) | 68 (97,1) | 25 (100) |  |
| Is there a cure for HPV? |  |  |  |  |  |
| Yes | 80 (69) | 50 (54,9) | 42 (60) | 13 (52,0) | 0,277 |
| No | 16 (13,8) | 25 (27,5) | 15 (21,4) | 6 (24,0) |  |
| Do not know | 20 (17,2) | 16 (17,6) | 13 (18,6) | 6 (24,0) |  |

¥: Chi-square; £: Fisher's Exact; HPV: Human papillomavirus.

The majority of individuals, both women (64.9%) and men (64.1%), reported knowing about the existence of the vaccine but were unable to say whether it was available at the nearest health unit (55.7% and 63.2%). More than 80% of individuals declared that they would vaccinate their children against HPV, but they were not vaccinated (86.5% and 92.3%). Many think that HPV infection has a cure (61.2%). Comparisons between genders showed no significant difference (p>0.05) for these questions, except for two. When asked what genital warts (better known as cockscomb) are, women had more knowledge (p=0.019) than men. The same occurred when asked if the child was vaccinated. Men, in general, did not know (p < 0.001).

### ASSOCIATION OF KNOWLEDGE ABOUT HPV IN QUILOMBOLAS IN DIFFERENT AGE GROUPS

When evaluating knowledge about HPV among the different generations of quilombolas, it is noted that the younger they are (18-34 years old), the better the knowledge. The majority of individuals aged between 18 and 34 had already heard of HPV (96.6%), compared to 48% of those over 65 years of age (p<0.001). Younger people (18-34 and 35-49 years old) claimed to know what HPV is, while older people (50-64 years old and over 65 years old) said they did not know (p<0.001). However, they all made the same mistake when reporting that HPV was a bacteria (p=0.155). The means of communication in which information about HPV was acquired varied according to the age group (p<0.001). The internet was the main source among individuals aged 18-34 years (26.7%), traditional means of communication (TV/foram/radio) were more prevalent in the age groups of 35-49 years (44%) and 50-64 years old (51.4%). People over 65 years reported not having heard of HPV anywhere (52%). Younger individuals (18 – 34 and 35 – 49 years old) knew about the existence of the vaccine, while the majority of those in the older age group (50 – 64 and > 65 years old) did not know (p<0.001). Despite this difference, regardless of age group, no one knew whether the vaccine was available at the nearest health unit (p=0.084). However, regardless of age group, the majority would vaccinate their children (p=0.250). The vaccination rate was higher in individuals between 18 and 34 years old (p<0.001). The 14 form of HPV transmission was better known among younger individuals (18 – 34 and 35 – 49 years old) (p<0.001), as well as whether the transmission was via sexual contact (p<0.029), and whether condoms were a form of prevention (p=0.008).

### ASSOCIATION OF KNOWLEDGE ABOUT HPV AND THE DIFFERENT QUILOMBOLAS AREAS IN THE CAXIAS REGION, MARANHÃO

Table 4 demonstrates a significant variation in sources of information about HPV between quilombola communities. There was a significant association (p = 0.004) concerning sources of information about HPV, highlighting that all communities, except Lavras, obtain information mainly through traditional media - TV/newspaper/radio. Lavras’s main source of HPV information comes from health professionals. Internet/Social media was the second source of HPV information in three of the six communities - Cajueiro, Jenipapo, and Mimoso. Significant disparities between communities existed regarding awareness about the availability of the HPV vaccine at the nearest healthcare facility. Jenipapo showed less knowledge, with only 6% of participants aware of the vaccine’s availability, while the majority (76.1%) were unaware of this information. Cajueiro had a more balanced distribution of knowledge, but most were unaware (60.0%) or didn’t know (27.5%) the information. Statistical analysis indicated a significant difference between communities in this aspect (p = 0.003). Mimoso community had the lowest proportion of affirmative responses regarding the vaccination of their children; only 16.7% declared that their children were vaccinated. In contrast, 41.2% of the Soledade community confirmed that their children had been vaccinated. The results, obtained using Fisher’s Exact Test, demonstrate a p-value of £0.045.

**Table 4.**
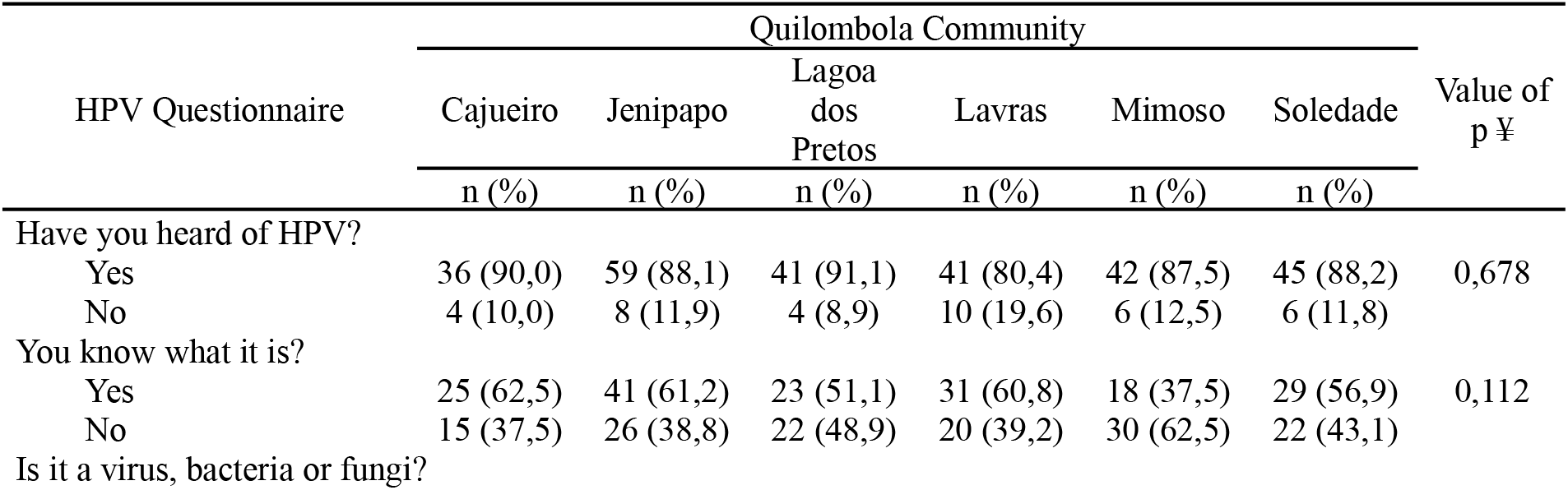
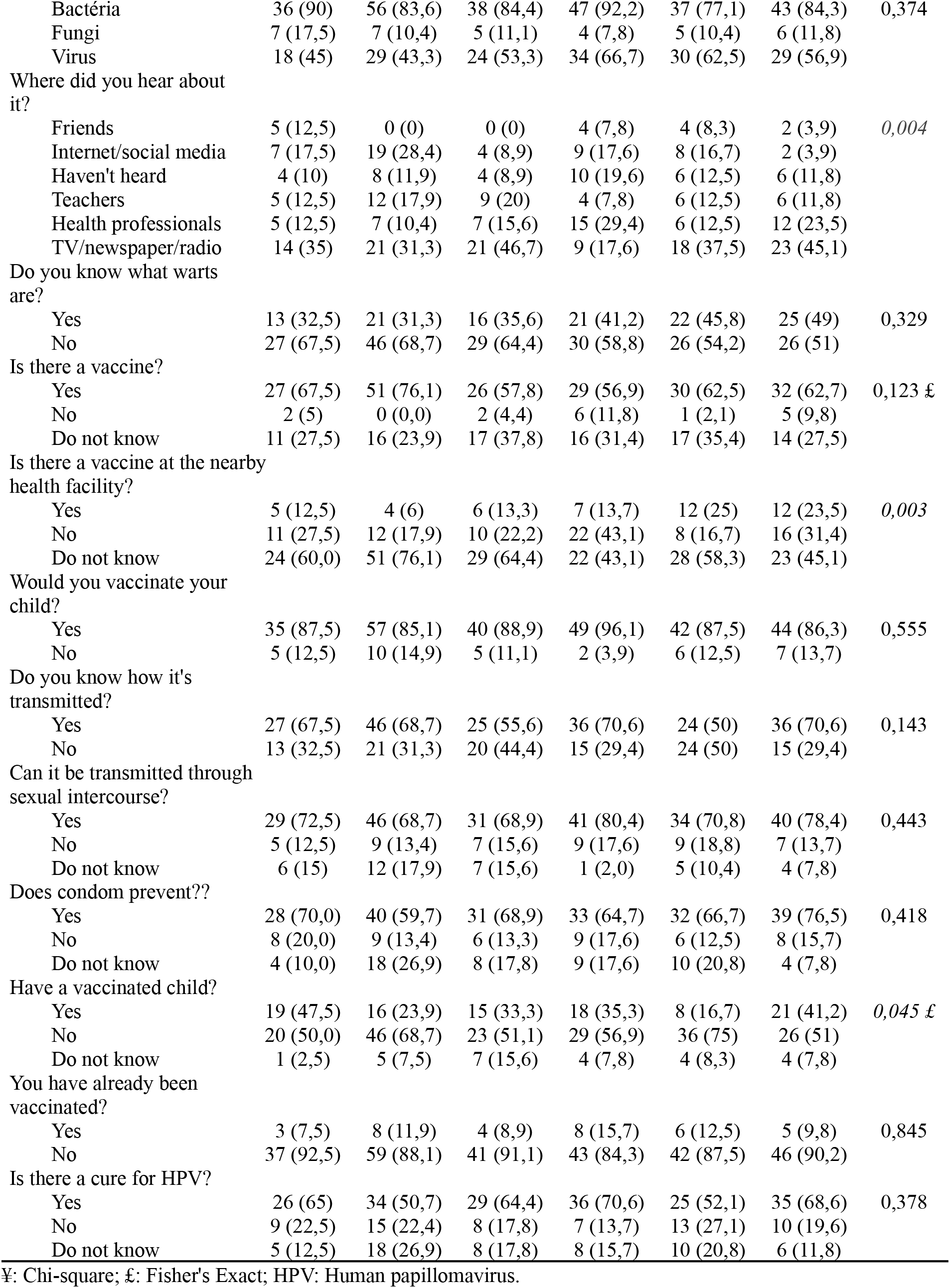
Knowledge and perceptions about HPV and its vaccination among different quilombola communities in Caxias, MA. Best time for São Luís 2023 in Maranhão, Brazil.

| HPV Questionnaire | Quilombola Community |  |  |  |  |  | Value of p ¥ |
| --- | --- | --- | --- | --- | --- | --- | --- |
|  | Cajueiro | Jenipapo | Lagoa dos Pretos | Lavras | Mimoso | Soledade |  |
|  | n (%) | n (%) | n (%) | n (%) | n (%) | n (%) |  |
| Have you heard of HPV? |  |  |  |  |  |  |  |
| Yes | 36 (90,0) | 59 (88,1) | 41 (91,1) | 41 (80,4) | 42 (87,5) | 45 (88,2) | 0,678 |
| No | 4 (10,0) | 8 (11,9) | 4 (8,9) | 10 (19,6) | 6 (12,5) | 6 (11,8) |  |
| You know what it is? |  |  |  |  |  |  |  |
| Yes | 25 (62,5) | 41 (61,2) | 23 (51,1) | 31 (60,8) | 18 (37,5) | 29 (56,9) | 0,112 |
| No | 15 (37,5) | 26 (38,8) | 22 (48,9) | 20 (39,2) | 30 (62,5) | 22 (43,1) |  |
| Is it a virus, bacteria or fungi? |  |  |  |  |  |  |  |
| Bactéria | 36 (90) | 56 (83,6) | 38 (84,4) | 47 (92,2) | 37 (77,1) | 43 (84,3) | 0,374 |
| Fungi | 7 (17,5) | 7 (10,4) | 5 (11,1) | 4 (7,8) | 5 (10,4) | 6 (11,8) |  |
| Virus | 18 (45) | 29 (43,3) | 24 (53,3) | 34 (66,7) | 30 (62,5) | 29 (56,9) |  |
| Where did you hear about it? |  |  |  |  |  |  |  |
| Friends | 5 (12,5) | 0 (0) | 0 (0) | 4 (7,8) | 4 (8,3) | 2 (3,9) | 0,004 |
| Internet/social media | 7 (17,5) | 19 (28,4) | 4 (8,9) | 9 (17,6) | 8 (16,7) | 2 (3,9) |  |
| Haven't heard | 4 (10) | 8 (11,9) | 4 (8,9) | 10 (19,6) | 6 (12,5) | 6 (11,8) |  |
| Teachers | 5 (12,5) | 12 (17,9) | 9 (20) | 4 (7,8) | 6 (12,5) | 6 (11,8) |  |
| Health professionals | 5 (12,5) | 7 (10,4) | 7 (15,6) | 15 (29,4) | 6 (12,5) | 12 (23,5) |  |
| TV/newspaper/radio | 14 (35) | 21 (31,3) | 21 (46,7) | 9 (17,6) | 18 (37,5) | 23 (45,1) |  |
| Do you know what warts are? |  |  |  |  |  |  |  |
| Yes | 13 (32,5) | 21 (31,3) | 16 (35,6) | 21 (41,2) | 22 (45,8) | 25 (49) | 0,329 |
| No | 27 (67,5) | 46 (68,7) | 29 (64,4) | 30 (58,8) | 26 (54,2) | 26 (51) |  |
| Is there a vaccine? |  |  |  |  |  |  |  |
| Yes | 27 (67,5) | 51 (76,1) | 26 (57,8) | 29 (56,9) | 30 (62,5) | 32 (62,7) | 0,123 £ |
| No | 2 (5) | 0 (0,0) | 2 (4,4) | 6 (11,8) | 1 (2,1) | 5 (9,8) |  |
| Do not know | 11 (27,5) | 16 (23,9) | 17 (37,8) | 16 (31,4) | 17 (35,4) | 14 (27,5) |  |
| Is there a vaccine at the nearby health facility? |  |  |  |  |  |  |  |
| Yes | 5 (12,5) | 4 (6) | 6 (13,3) | 7 (13,7) | 12 (25) | 12 (23,5) | 0,003 |
| No | 11 (27,5) | 12 (17,9) | 10 (22,2) | 22 (43,1) | 8 (16,7) | 16 (31,4) |  |
| Do not know | 24 (60,0) | 51 (76,1) | 29 (64,4) | 22 (43,1) | 28 (58,3) | 23 (45,1) |  |
| Would you vaccinate your child? |  |  |  |  |  |  |  |
| Yes | 35 (87,5) | 57 (85,1) | 40 (88,9) | 49 (96,1) | 42 (87,5) | 44 (86,3) | 0,555 |
| No | 5 (12,5) | 10 (14,9) | 5 (11,1) | 2 (3,9) | 6 (12,5) | 7 (13,7) |  |
| Do you know how it's transmitted? |  |  |  |  |  |  |  |
| Yes | 27 (67,5) | 46 (68,7) | 25 (55,6) | 36 (70,6) | 24 (50) | 36 (70,6) | 0,143 |
| No | 13 (32,5) | 21 (31,3) | 20 (44,4) | 15 (29,4) | 24 (50) | 15 (29,4) |  |
| Can it be transmitted through sexual intercourse? |  |  |  |  |  |  |  |
| Yes | 29 (72,5) | 46 (68,7) | 31 (68,9) | 41 (80,4) | 34 (70,8) | 40 (78,4) | 0,443 |
| No | 5 (12,5) | 9 (13,4) | 7 (15,6) | 9 (17,6) | 9 (18,8) | 7 (13,7) |  |
| Do not know | 6 (15) | 12 (17,9) | 7 (15,6) | 1 (2,0) | 5 (10,4) | 4 (7,8) |  |
| Does condom prevent?? |  |  |  |  |  |  |  |
| Yes | 28 (70,0) | 40 (59,7) | 31 (68,9) | 33 (64,7) | 32 (66,7) | 39 (76,5) | 0,418 |
| No | 8 (20,0) | 9 (13,4) | 6 (13,3) | 9 (17,6) | 6 (12,5) | 8 (15,7) |  |
| Do not know | 4 (10,0) | 18 (26,9) | 8 (17,8) | 9 (17,6) | 10 (20,8) | 4 (7,8) |  |
| Have a vaccinated child? |  |  |  |  |  |  |  |
| Yes | 19 (47,5) | 16 (23,9) | 15 (33,3) | 18 (35,3) | 8 (16,7) | 21 (41,2) | 0,045 £ |
| No | 20 (50,0) | 46 (68,7) | 23 (51,1) | 29 (56,9) | 36 (75) | 26 (51) |  |
| Do not know | 1 (2,5) | 5 (7,5) | 7 (15,6) | 4 (7,8) | 4 (8,3) | 4 (7,8) |  |
| You have already been vaccinated? |  |  |  |  |  |  |  |
| Yes | 3 (7,5) | 8 (11,9) | 4 (8,9) | 8 (15,7) | 6 (12,5) | 5 (9,8) | 0,845 |
| No | 37 (92,5) | 59 (88,1) | 41 (91,1) | 43 (84,3) | 42 (87,5) | 46 (90,2) |  |
| Is there a cure for HPV? |  |  |  |  |  |  |  |
| Yes | 26 (65) | 34 (50,7) | 29 (64,4) | 36 (70,6) | 25 (52,1) | 35 (68,6) | 0,378 |
| No | 9 (22,5) | 15 (22,4) | 8 (17,8) | 7 (13,7) | 13 (27,1) | 10 (19,6) |  |
| Do not know | 5 (12,5) | 18 (26,9) | 8 (17,8) | 8 (15,7) | 10 (20,8) | 6 (11,8) |  |

¥: Chi-square; £: Fisher's Exact; HPV: Human papillomavirus.

## DISCUSSION

Despite the advent of vaccines, prevention, and screening recommendations, cervical cancer still ranks as the fourth most common type among women in the world, with 660.000 new cases per year and responsible for 350.000 deaths (Ferlay et al., 2021). In Brazil, it is the third most common type of cancer and the fourth cause of death from cancer among women – especially black, poor women, and women with low levels of formal education (Pan American Health Organization, 2023). Cervical cancer is highly preventable and can be easily treated if detected at an early stage. However, there is a disproportionately high impact on cervical cancer incidence and mortality in low- and middle-income countries that do not have organized screening and prevention programs (Gallagher et al., 2018). Cost-effective strategies and tools are needed to reduce the impact of cervical cancer worldwide to mitigate existing disparities in cervical cancer rates between resource-poor and resource-rich settings (Pimple, 2019). This study represents one of the first studies in Brazil to investigate knowledge about HPV and its vaccination, aimed at quilombola men and women. The literature related to HPV and quilombola communities in Brazil focuses on women, especially on the association of HPV infection with cervical cancer (Figueredo et al., 2022; Nascimento et al., 2017; de Ribamar Ross et al., 2023). The emphasis on women’s health, although crucial, leaves gaps in knowledge and awareness about HPV among the male quilombola population. A more equitable approach is extremely important to promote the overall health of the quilombola community and mitigate potential gender disparities in perception and access to HPV-related information.

Our methodology differed from other similar studies around the world that were carried out with traditional communities. Alsous and collaborators (2021) assessed the knowledge and awareness of women from Arab communities in 4 Middle Eastern countries about HPV, infection, and vaccination, and the questionnaire was distributed using the Google Forms electronic format, reaching a greater number of women (n=3658) (Alsous et al., 2021). However, the authors themselves report the possibility of selection bias due to the online nature of the survey and the possibility of careless responses either due to needing to understand the question or due to the rush to finish the questionnaire or looking up answers online. For quilombola communities, this methodology would not work since internet access is limited due to the quality of the service or because individuals need computers or other equipment to answer an online questionnaire. Furthermore, due to the low level of education observed in the research, many quilombola would need an adequate understanding or wouldn’t be able to read the questions.

In the research by Raçi and collaborators (2021) on women from Kosovo (n=645), a low-income country located in Eastern Europe, the questionnaire was given to women and self-answered (Raçi et al., 2021). The low educational level of quilombolas can lead to a misunderstanding of what is written in the questionnaire, leading to null or careless answers. Carrying out the questionnaires in person reduces these biases; however, as the face-to-face application of the questionnaires is more time-consuming and laborious, the range of individuals ends up being smaller.

It is clear that although the majority of participants have already heard about HPV, a large proportion are still unaware of the type of microorganism that HPV represents and presents a noted confusion between HPV and HIV. This confusion highlights the importance of clarifying the differences between these infections to ensure accurate and informed understanding, as this confusion can lead to prejudice in seeking help and health services. The study conducted by Dias in 2021, which focuses on the prevalence of STIs in quilombola women in Brazil, highlights the pressing need to increase awareness and access to health services (Dias et al., 2021). The results indicate a clear correlation between lack of awareness and a higher incidence of STIs, including HPV. The presence of healthcare facilities and knowledge about vaccines, especially the HPV vaccine, play an extremely important role in the health disparities present in specific communities.

In our study, traditional media (TV, newspapers, radio) proved to be the main source of information about HPV, especially among older age groups. This was also seen by Katz et al. (2011) and Rosen et al. (2017), who, although investigated a different population from ours, identified similar results, highlighting TV or TV advertisements as one of the main sources of information about HPV and its vaccination (Katz et al., 2011; Rosen et al., 2017). Although the internet is a more significant source for younger people, it still takes a backseat in older age groups. A study that examined the relationship between the use of social media and knowledge about HPV showed results that indicate that the use of these platforms is associated with greater awareness about HPV and its vaccine, suggesting that strategies based on social media can be effective in promoting knowledge about HPV and encouraging vaccine uptake, potentially reducing cancer incidence rates in vulnerable populations. The increasing access to the internet, especially among the younger population, highlights the great potential of social networks as a crucial tool in disseminating health information (Katz et al., 2011; Rosen et al., 2017).

Chandeying and colleagues (2023) conducted a systematic review and meta-analysis demonstrating the importance of digital interventions to promote HPV vaccination (Chandeying; Thongseiratch, 2023). The authors emphasise using technology and online platforms to resolve information barriers about the virus, provide education about vaccination, clarify myths, and send reminders and alerts to families about vaccination appointments. Fadia Dib and colleagues (2021) demonstrated that approximately 24% of the interviewed women (466/1949) reported using the Internet as a source of vaccine information (Vonasek et al., 2016).

In our work, individuals aged 18 to 34 (38.4%) used the Internet as the most common means of obtaining information about HPV. However, this was not observed among older people who still use TV/Radio/Newspapers to obtain information.

The comparative analysis between genders revealed significant differences in knowledge about genital warts, with women demonstrating a more comprehensive understanding. The lack of information among men about their children’s vaccination status highlights possible disparities in perception and access to information within quilombola communities, suggesting a pattern in which responsibility for childcare often falls primarily to mothers. This observation echoes a study by Vonasek et al. (2016), which revealed that 93.5% of Uganda’s women interviewed understood that childhood immunisations protect children from diseases (Rosen et al., 2017). The same was observed in Osazuwa-Peters et al. (2017), where it was noted that men and women in the United States were more likely to have never heard about HPV or the vaccine compared with women (Fadia et al., 2021). This study’s results highlighted mothers’ central role in vaccinating their children, as they not only seek information and discuss the issue with health professionals but also have the power to decide which vaccines are administered. Women’s knowledge about their children’s vaccination status in the quilombola communities suggests that mothers are more active and involved in caring for and protecting their children’s health.

The data points to an increase in knowledge as the age range decreases, indicating a significant correlation between youth and understanding of HPV. Among younger quilombolas (18-34 years old), the majority have heard of HPV compared to older ones (over 65 years old). One hypothesis that can be raised to explain this phenomenon is the focus of health education campaigns on HPV. In the Brazilian context, the recommended age range for HPV vaccination is 9 to 14 years old, with exceptions to this recommendation. Guidelines for HPV vaccination may vary for immunocompromised people, whose protection needs may require different strategies. Furthermore, more recently, recommendations have been expanded to include people who have experienced sexual violence, recognizing the importance of vaccination in these cases to prevent transmission of the virus.

A non-random relationship between the “quilombo of residence” and the sources of information to obtain knowledge about HPV opens space for exploring some hypotheses. For example, communities in extremely rural areas (Figure 1 Supplementary material), such as Lavras, Cajueiro, and Mimoso, may face difficulties accessing information about HPV, possibly due to the lack of availability and accessibility of health services. The Same was shown by Mohammed and colleagues (2018) when analysing the knowledge among residents of rural areas compared to urban areas in the United States; it was observed that rural residents were less likely to know what HPV is, its vaccine, or cancers associated with its infection (Osazuwa-Peters et al., 2017). Besides the differences between population targets, this variety in knowledge about HPV and inaccessibility to different sources of information highlights the need to consider not only the community itself but also the geographic location in evaluating and implementing possible strategies to improve the knowledge of this population.

**Figure 1.**
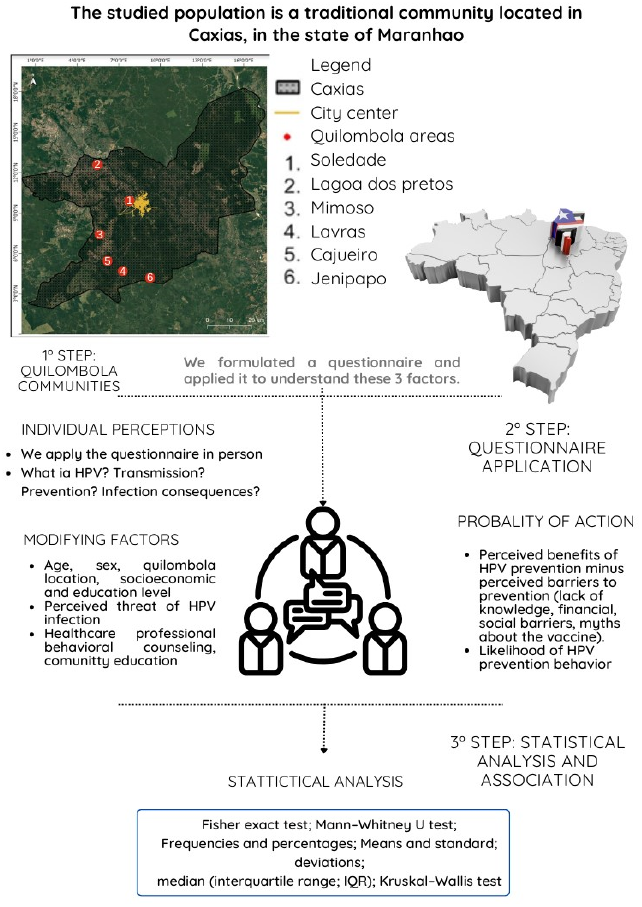
Locations map of quilombola communities and experimental design of methodology.

Another hypothesis to be explored is the presence of health units in the communities or proximity to them. Quilombola communities that have local health units can have a direct source of health information, including HPV, which positively influences knowledge and perception about it. The correlation between the availability of health facilities and the information received can be a significant factor in understanding the disparities observed in community responses. Considering the study by Mota et al. (2021), which presents data on the lack of access to health in certain quilombola groups and expands the scope of the discussion to general access to health in quilombola populations in Brazil, we can visualise the complexity of disparities in access to health and in obtaining information about specific health issues, such as HPV, highlighting the need for specific strategies and approaches to improve the situation of quilombola populations in the public health context (Lama et al., 2021).

The Jenipapo community’s low awareness rate about vaccine availability may reflect a lack of direct access to health services. In contrast, the Cajueiro community’s more balanced distribution of knowledge may indicate better integration with local health services. The statistical analysis reinforces the relevance of these hypotheses, with a significant p (p = 0.003), suggesting that factors such as geographic location and the presence of health units impact the knowledge and perception of HPV among the population in the study communities.

Our data depict the situation of a traditional community and its understanding of HPV and its vaccine. Our work and hypotheses underscore the necessity for further research concerning quilombola communities, as well as new dissemination strategies regarding HPV vaccination within these communities. Our findings and efforts address a gap in the literature concerning traditional communities, particularly quilombolas, and contribute gradually to elucidating the extent of change and improvement needed for public health among vulnerable populations in Maranhão, Brazil.

## Data Availability

All data produced in the present study are available upon reasonable request to the authors

## Supplementary Information

The online version contains supplementary material available

## Author Contribution

FV designed the study, MC prepared the first draft, and LC worked on the subsequent versions of the manuscript under supervision and collaboration with FV. JR, RM, and GC collected all the data, and FV, LV, and LC analysed it. NE and AC reviewed and edited previous versions of the manuscript. All authors read and approved the final manuscript.

## Data Availability

The focus group data are not publicly available. The data belong to the Quilombola community and the Federal University of Maranhão.

## Declarations Ethical Statement

The study is approved by the Research Ethics Committee of the Fundação Universidade Federal do Maranhão, number: 5.836.549.

## Acknowledgment

We would like to thank Professor Ross’ entire nursing team at the State University of Maranhão, Caxias campus, for their collaboration and help in this work, and the whole Brazilian quilombola population for their daily struggle for better living conditions. The Foundation for Supporting Research and Scientific and Technological Development of Maranhão (FAPEMA) supports the research (Grant INFRA02003/21).

## Footnotes

1 The word “quilombo” originates from the term “kilombo,” present in the language of the Bantu peoples, originating from Angola, and it means a place of rest or encampment. “Quilombola” is used for traditional communities in Brazil, composed of descendants and remnants of communities formed by runaway slaves during the period of slavery (1500 to 1888). These are geographically isolated communities with their own customs and knowledge passed down through generations. Currently, these communities face difficulties in accessing healthcare and education

## References

[1] Doorbar J, Quint W, Banks L, Bravo IG, Stoler M, Broker TR, Stanley MA. The biology and life-cycle of human papillomaviruses. Vaccine. 2012 Nov 20;30 Suppl 5:F55–70. doi: 10.1016/j.vaccine.2012.06.083. PMID: 23199966.

[2] World health statistics 2023: monitoring health for the SDGs, Sustainable Development Goals. Geneva: World Health Organization;2023. Licence: CC BY-NC-SA 3.0 IGO.

[3] García, F.R., Norenhag, J., Edfeldt, G. et al. Prevalence of the human papillomavirus (HPV) types among cervical dysplasia women attending a gynaecological clinic in Sweden. BJC Rep 1, 11 (2023). 10.1038/s44276-023-00012-y

[4] Alsous MM, Ali AA, Al-Azzam SI, Abdel Jalil MH, Al-Obaidi HJ, Al-Abbadi EI, Hussain ZK, Jirjees FJ. Knowledge and awareness about human papillomavirus infection and its vaccination among women in Arab communities. Sci Rep. 2021 Jan 12;11(1):786. doi: 10.1038/s41598-020-80834-9. PMID: 33436959; PMCID: PMC7804285.

[5] Chandeying, N., Thongseiratch, T. Systematic review and meta-analysis comparing educational and reminder digital interventions for promoting HPV vaccination uptake. npj Digit. Med. 6, 162 (2023). 10.1038/s41746-023-00912-w

[6] Waller J, Ostini R, Marlow LA, McCaffery K, Zimet G. Validation of a measure of knowledge about human papillomavirus (HPV) using item response theory and classical test theory. Prev Med. 2013 Jan;56(1):35–40. doi: 10.1016/j.ypmed.2012.10.028. Epub 2012 Nov 8. PMID: 23142106.

[7] Gallagher KE, LaMontagne DS, Watson-Jones D. Status of HPV vaccine introduction and barriers to country uptake. Vaccine. 2018 Aug 6;36(32 Pt A):4761–4767. doi: 10.1016/j.vaccine.2018.02.003. Epub 2018 Mar 23. PMID: 29580641.

[8] Raçi PZ, Raçi F, Hadri T. Kosovo women’s knowledge and awareness of human papillomavirus (HPV) infection, HPV vaccination, and its relation to cervical cancer. BMC Womens Health. 2021 Oct 9;21(1):354. doi: 10.1186/s12905-021-01496-x. PMID: 34627223; PMCID: PMC8502331.

[9] Warner ZC, Reid B, Auguste P, Joseph W, Kepka D, Warner EL. Awareness and Knowledge of HPV, HPV Vaccination, and Cervical Cancer among an Indigenous Caribbean Community. Int J Environ Res Public Health. 2022 May 7;19(9):5694. doi: 10.3390/ijerph19095694. PMID: 35565089; PMCID: PMC9105034.

[10] de Ribamar Ross, J., Marinelli, N.P., Vidal, F.C.B. et al. Frequency of human papilomavirus and associated factors in gypsy and quilombola women. BMC Women’s Health 23, 160 (2023). 10.1186/s12905-023-02239-w

[11] Dias JA, Luciano TV, Santos MCLFS, Musso C, Zandonade E, Spano LC, Miranda AE. Infecções sexualmente transmissíveis em mulheres afrodescendentes de comunidades quilombolas no Brasil: prevalência e fatores associados. Cadernos de Saúde Pública 2021; 37(2):e00174919 doi: 10.1590/0102-311x00174919

[12] IBGE – INSTITUTO BRASILEIRO DE GEOGRAFIA E ESTATíSTICA. Censo Demográfico 2022 Quilombolas. Primeiros resultados do universo. Rio de Janeiro: IBGE, 2023.

[13] Lima, L. B.; Melo, A. F. de.; Barbosa, D. R. e S. The quilombola territory in communities in the interior of Brazilian Northeast: preliminary socioeconomic and structural characterization. Research, Society and Development, [S. l.], v. 10, n. 13, p. e452101320899, 2021. DOI: 10.33448/rsd-v10i13.20899.

[14] Ferlay J, Ervik M, Lam F, Colombet M, Mery L, Piñeros M, et al. Global Cancer Observatory: Cancer Today. Lyon: International Agency for Research on Cancer; 2020 (https://gco.iarc.fr/today, accessed February 2024).

[15] Pan American Health Organization. Risk Country Cooperation Strategy 2022-2027 Brazil. Revised Version. Brasilia, D.F.: PAHO; 2023.

[16] Pimple SA, Mishra GA. Global strategies for cervical cancer prevention and screening. Minerva Ginecol 2019;71:313–20. DOI: 10.23736/S0026-4784.19.04397-1

[17] Figueredo, R. L., Durans, K. C. N., Abreu, C. M. R., Batista, M. C. A., Barros, L. A. A., Morais, C.D.M.de, & Freitas, D. da S. (2022). Knowledge profile of quilombola women about uterine cervical cancer in the village of Santana dos pretos. Brazilian Journal of Development, 8(2), 13081–13097. 10.34117/bjdv8n2-307

[18] Nascimento VB, Martins NVN, Ciosak SI et al. Vulnerabilidades de mulheres quilombolas no interior da Amazônia às infecções sexualmente transmissíveis: um relato de experiência. Interdisciplinary Journal of Health Education. 2017 Jan-Jul;2(1):68–73. 10.4322/ijhe.2016.029

[19] Katz ML, Krieger JL, Roberto AJ. Human papillomavirus (HPV): college male’s knowledge, perceived risk, sources of information, vaccine barriers and communication. J Mens Health. 2011 Oct 1;8(3):175–184. doi: 10.1016/j.jomh.2011.04.002.

[20] Rosen BL, Shew ML, Zimet GD, Ding L, Mullins TLK, Kahn JA. Human Papillomavirus Vaccine Sources of Information and Adolescents’ Knowledge and Perceptions. Glob Pediatr Health. 2017 Nov 24;4:2333794X17743405. doi: 10.1177/2333794X17743405.

[21] Lama, Y., Quinn, S. C., Nan, X., & Cruz-Cano, R. (2021). Social media use and human papillomavirus awareness and knowledge among adults with children in the household: examining the role of race, ethnicity, and gender. Human Vaccines & Immunotherapeutics, 17(4), 1014–1024. 10.1080/21645515.2020.1824498

[22] Vonasek BJ, Bajunirwe F, Jacobson LE, Twesigye L, Dahm J, Grant MJ, Sethi AK, Conway JH. Do Maternal Knowledge and Attitudes towards Childhood Immunizations in Rural Uganda Correlate with Complete Childhood Vaccination? PLoS One. 2016 Feb 26;11(2):e0150131. doi: 10.1371/journal.pone.0150131. PMID: 26918890; PMCID: PMC4769080.

[23] Fadia Dib, Philippe Mayaud, Laetitia Longfier, Pierre Chauvin, Odile Launay, Effect of Internet use for searching information on vaccination on the uptake of human papillomavirus vaccine in France: A path-analysis approach, Preventive Medicine, Volume 149, 2021, 106615, ISSN 0091-7435, 10.1016/j.ypmed.2021.106615.

[24] Osazuwa-Peters N, Adjei Boakye E, Mohammed KA, Tobo BB, Geneus CJ, Schootman M. Not just a woman’s business! Understanding men’s and women’s knowledge of HPV, the HPV vaccine, and HPV-associated cancers. Prev Med. 2017 Jun;99:299–304. doi: 10.1016/j.ypmed.2017.03.014. Epub 2017 Mar 22. PMID: 28341458.

[25] Mohammed KA, Subramaniam DS, Geneus CJ, Henderson ER, Dean CA, Subramaniam DP, Burroughs TE. Rural-urban differences in human papillomavirus knowledge and awareness among US adults. Prev Med. 2018 Apr;109:39–43. doi: 10.1016/j.ypmed.2018.01.016. Epub 2018 Jan 31. PMID: 29378268.

[26] Mota AN, Maciel ES, Quaresma FRP, de Araújo FA, Sousa LVA, Junior HM, Fonseca FLA, Adami F. A look at vulnerability: analysis of the lack of access to health care for quilombolas in Brazil. J Hum Growth Dev. 2021; 31(2):302–309. DOI: 10.36311/jhgd.v31.11404

